# Reduced option generation reveals distinct profiles of apathy in schizophrenia

**DOI:** 10.64898/2026.08.03.26359552

**Authors:** Noham Wolpe, Cheng-Lin Wu, Clàudia Aymerich, Marta Martin-Subero, Paloma Fuentes-Perez, Claudia Ovejas-Catalan, Renata Zirilli, Sophie Shatford, Rebecca Cox, Megan Cartier, Ana Catalan, Anna Mane, John Pratt, Lisa Airey, Javier Vazquez-Bourgon, Nuria Segarra, Yi-Jie Zhao, Paul C Fletcher, Peter B Jones, Masud Husain, Emilio Fernandez-Egea

**Author notes:** Corresponding author: Emilio Fernandez-Egea MD PhD Department of Psychiatry University of Cambridge, 128 Tenison road, CB1 2DP Cambridge, UK.

## Abstract

Apathy is a major driver of long-term disability in schizophrenia, yet existing accounts focus on how patients evaluate the cost of action while neglecting their capacity to generate options for action in the first place. Using a brief, culture-fair option generation task (OGT) within the international CHANSS study, we examined this process in 150 patients with different stages of schizophrenia and 100 healthy controls across the UK, Spain, and China. The OGT requires drawing as many different paths as possible between two points on a touchscreen, yielding two measures: fluency (the number of paths that they generate) and uniqueness (the distinctiveness of those paths). Patients drew as many paths as controls but produced options roughly half as unique. Across patients, uniqueness was positively associated with verbal fluency and depressive symptom severity, but negatively associated with self-reported behavioural apathy. This suggests that increased apathy is associated with reduced ability to generate unique options (paths on the task).

To examine the cognitive mechanisms further, we decomposed uniqueness into four exploratory measures: roaming entropy (spatial breadth of exploration), option elaboration (within-path development), perseveration (local repetition), and novelty maintenance (sustaining diversity over time). Together these explained 69% of the variance in motor-residualised uniqueness and attenuated the group difference by 80%, with roaming entropy as the dominant contributor. K-means clustering revealed three distinct patient profiles, each with a different clinical signature. A small subgroup generated options much like the control group. A larger subgroup retained spatial exploration but developed each option less and reported more depressive symptoms. A third subgroup showed broadly impaired exploration and the lowest verbal fluency. These findings establish option generation as a measurable behavioural phenotype in schizophrenia that relates to apathy and can be decomposed into specific cognitive mechanisms. They suggest that an impoverished option space, marked by reduced generation of unique possibilities for action, may be a remediable component of apathy.

## Introduction

Schizophrenia is a chronic and disabling psychiatric disorder affecting approximately 1% of the global population and representing a leading cause of disability worldwide ^1^. Although considerable research has focused on the pathophysiology of positive symptoms, negative symptoms account for a disproportionate share of long-term functional impairment and remain poorly responsive to current pharmacological and psychosocial interventions ^2,3^. Negative symptoms are conventionally organised into two principal dimensions: diminished emotional expression (blunted affect and alogia) and deficits in motivation and pleasure (avolition, anhedonia, and asociality) ^4–6^. Emerging evidence indicates that these dimensions are psychometrically and potentially biologically ^7–9^ and therapeutically ^10^ distinct. Despite their clinical importance, motivational impairment, operationally subsumed here under the term ‘apathy’, referring to a reduction in goal-directed behaviour, remains poorly characterised cognitively and correspondingly difficult to treat^4^.

A growing body of research has approached motivational deficits in schizophrenia through the lens of decision-making, and particularly effort-based decision-making^11–13^. This line of research has demonstrated that patients are less willing to exert physical or cognitive effort in exchange for a reward^14–16^. However, effort-based decision-making paradigms require the experimenter to specify the options from which participants choose, bypassing the prior process of generating those options in the first place. In everyday goal-directed behaviour, ‘option generation’, or the ability to self-generate a repertoire of possible actions,, is a critical precondition for subsequent evaluation and selection^17,18^.

Despite its theoretical importance, option generation has rarely been studied in psychiatric populations. In schizophrenia, one study found reduced capacity to generate options verbally for ill-structured scenarios, but the task relied on verbal production, which is itself impaired in schizophrenia and was partly implicated in the observed association with apathy^19^. A nonverbal task is therefore needed to test option generation in schizophrenia. The conceptual framework linking executive cognition, motivational cognition, and meta-cognition to motivational impairment in schizophrenia positions option generation as a core component of executive cognition within this syndrome^20^. The ability to plan, initiate, and sustain goal-directed actions depends in part on the capacity to generate a sufficient and diverse repertoire of possible courses of action^17^. Executive dysfunction, and particularly impaired verbal fluency, the ability to generate different words of the same letter or category, is well established in schizophrenia and is consistently linked to negative symptoms^21–23^, providing a plausible link to option generation deficits.

The option generation task (OGT) provides a quantitative, objective, and culture-fair measure of this process^17^. Participants draw as many different paths as possible between two fixed points on a touchscreen within a defined time window. Two main metrics are extracted. The first is ***fluency*** which is the total number of paths generated. The second measure is ***uniqueness***, which is the geometric dissimilarity of each path relative to all other paths in the dataset. In other words, this a metric of how unique each path is relative to the others drawn. In healthy individuals, there is a trade-off between these dimensions: people tend to produce either many similar options or fewer, more distinctive ones^17,24^. Crucially, adding a motor control condition lets the OGT isolate the contribution of basic motor features. Uniqueness can then be interpreted as a cognitive capacity rather than a movement one. Unlike many tasks, including verbal fluency, it is also independent of education level, which can otherwise confound performance.

Two bodies of prior work motivate the study of option generation in schizophrenia. First, dopamine replacement therapy in Parkinson’s disease shifts behaviour along the fluency-uniqueness spectrum. In this disorder, higher dopaminergic tone increased fluency (number of paths generated) but reduced their uniqueness. Crucially, however, even after correcting for this trade-off, dopamine improved uniqueness for a given fluency level^17^. Second, patients with major depressive disorder (MDD) generate fewer options than healthy controls, but those options are more unique^24^. This pattern complicates a simple motivational account of option generation, because in healthy volunteers, greater motivation is associated with generating *more* options of *lower* uniqueness^17^. In a subset of these MDD patients, PET imaging showed that putamen D2/D3 receptor availability correlated negatively with fluency and positively with uniqueness. This suggests that reduced dopaminergic occupancy in the putamen drives the generation of fewer, more idiosyncratic options in depression. Together, these data in Parkinson’s disease and MDD implicate dopaminergic modulation of fronto-striatal circuits in option generation and raise the question of how this process is affected in schizophrenia, where both dopaminergic dysregulation and executive dysfunction are prominent features.

The present study had two primary aims. First, we examined whether patients with schizophrenia, tested as part of a multisite study CHANSS^20^, show impaired option generation relative to healthy controls, and characterised the direction and specificity of any deficit. Second, we investigated the clinical correlates of option generation performance within the patient group, asking which specific dimensions of cognition and apathy explain individual differences in uniqueness and whether it is related to depressive symptoms. We further decomposed the behavioural differences in uniqueness to identify more specifically the cognitive mechanisms that drive the group-level differences. We used these mechanisms to stratify the patient group through a clustering method, revealing distinct behavioural profiles in option generation and their clinical profiles.

## Methods

### Study design and participants

This study used data from CHANSS (Characterising Negative Symptoms in Schizophrenia), a multicentre, experimental, cross-sectional study designed to investigate the cognitive mechanisms underlying motivational impairment in schizophrenia^20^. As described in the CHANSS protocol, recruitment spanned multiple sites in the UK, Spain and China and targeted a clinically heterogeneous sample across illness stages, including patients with early psychosis (within 5 years of first episode) and treatment-resistant schizophrenia. The protocol specified inclusion criteria of age 18–65 years, ICD-10 schizophrenia diagnosis of at least 1-year, stable antipsychotic treatment for at least 6 weeks, and capacity to provide informed consent. Exclusion criteria included neurological or major medical disorders affecting cognition, traumatic brain injury with loss of consciousness greater than 5 min, current substance use disorder other than nicotine, neurodevelopmental disorder, intellectual disability (IQ < 70), and anticholinergic medication use other than hyoscine for clozapine-induced sialorrhoea. The study was approved by the relevant local research ethics committees at each site, and all participants provided written informed consent. Control participants were tested at Tel Aviv University (ethics ref: 0005906). All participants provided informed consent before starting the experiment. The dataset comprised 250 participants: 150 patients from the CHANSS study and 100 healthy controls (Table 1).

**Table 1.** Values are mean (SD), median [IQR], or n (%). Inferential 1 tests are Welch t-tests for continuous measures and chi-square tests for categorical measures; EP = early psychosis within years of illness onset; TRS = treatment-resistant schizophrenia, prescribed with clozapine; Antipsychotic load expressed as % Maximum British National Formulary; AMI = Apathy Motivation Index; BACS = Brief Assessment of Cognition in Schizophrenia; BNSS MAP/EXP = Brief Negative Symptom Scale, Motivation and Please/ Expressive deficit subscales; CDSS = Calgary Depression Scale for Schizophrenia; PSP = Personal and Social Performance scale.

|  | Patients (n = 150) | Controls (n = 100) | Inferential statistics |
| --- | --- | --- | --- |
| **Demographic** |  |  |  |
| Age, years | 42.5 (10.3), n = 149 | 25.7 (4.2), n = 89 | $t = 17.60, p < .001$ |
| Sex, female / male | 63 / 87, n = 150 | 70 / 19, n = 89 | $\chi^2 = 30.40, p < .001$ |
| Handedness, right / left | -- | 75 / 14, n = 89 |  |
| Education, years | 14.6 (3.7), n = 145 | 15.1 (2.3), n = 89 | $t = -1.24, p = 0.22$ |
| **Clinical** |  |  |  |
| Site, UK / Spain / China | 87 / 28 / 35, n = 150 |  |  |
| Age at first episode, years | 25.1 (7.8), n = 149 |  |  |
| Illness duration, years | 17.4 (10.5), n = 148 |  |  |
| EP | 31 (20.9%), n = 148 |  |  |
| TRS | 53 (35.3%), n = 150 |  |  |
| Antipsychotic burden | 77.9% (48.2%), n = 150 |  |  |
| Antipsychotic polypharmacy | 54, n = 150 |  |  |
| Current antidepressant prescription | 51, n = 150 |  |  |
| **Cognitive** |  |  |  |
| BACS composite z-score | -1.5 (1.3), n = 149 |  |  |
| BACS verbal fluency z-score | -1.2 (1.2), n = 149 |  |  |
| BACS digit sequencing z-score | -1.0 (1.7), n = 149 |  |  |
| BACS trail-making z-score | -0.6 (1.4), n = 149 |  |  |
| **Clinical** |  |  |  |
| PANSS Marder positive | 6.7 (3.2), n = 150 |  |  |
| BNSS MAP | 10.0 (7.4), n = 150 |  |  |
| BNSS EXP | 5.9 (5.7), n = 150 |  |  |
| AMI behavioural | 2.3 (0.8), n = 140 |  |  |
| AMI social | 2.1 (0.8), n = 140 |  |  |
| AMI emotional | 2.6 (0.7), n = 140 |  |  |
| CDSS total | 3.6 (4.5), n = 150 |  |  |
| PSP score | 67.0 (15.6), n = 150 |  |  |

### CHANSS assessment battery and clinical measures

The CHANSS protocol combined structured clinical ratings, self-report questionnaires, standard neurocognitive assessment and four computerised tasks probing executive cognition, motivational cognition and meta-cognition^20^. For the present analyses, we focused on the option generation task and a prespecified subset of clinical measures available in the CHANSS clinical database. These included Brief Assessment of Cognition in Schizophrenia (BACS) subdomains^25^ of verbal memory, digit sequencing, token motor, verbal fluency, symbol coding and Tower of London; Personal and Social Performance (PSP)^26^; Calgary Depression Scale for Schizophrenia (CDS)^27^; and Apathy Motivation Index (AMI) behavioural, social and emotional subscales, computed according to the original scoring^28^. We additionally derived motivational and expressive negative symptom dimensions from the Brief Negative Symptom Scale (BNSS)^29^. A motivation and pleasure score (MAP) was computed as the sum of BNSS items 1, 2, 3, 5, 6, 7 and 8, and an emotional expressivity score (EXP) as the sum of BNSS items 9, 10, 11, 12 and 13. PANSS Marder positive symptom scores were computed from items P1, P3, G9 and G10^30^.

### Option generation task

The option generation task is an assay of cognitive flexibility, action fluency and creative option generation^17^. Participants performed the task seated in front of a touchscreen tablet using a stylus (Fig. 1). The task was conducted on a Samsung Galaxy Tab S6 Lite tablet. Two red circles were shown on the screen, one at the bottom and one at the top. In the initial control condition, participants were instructed to draw as many paths as possible between the two circles in 30 s. In the main test condition, they were instructed to draw as many different paths as possible between the same circles in 120 s. Paths remained visible on the screen to reduce working memory demands. Paths could be straight or curved and could overlap or intersect, but to be counted as valid they had to begin at the lower circle and terminate at the upper circle. The control condition was intended to account for inter-individual differences in basic drawing speed, while the test condition yielded both fluency measure (number of options generated) and the main uniqueness measure described below. Due to a technical error in the control condition, data for the control condition was missing for two patients, leaving 148 patients for analyses involving control-condition motor measures.

**Figure 1.**
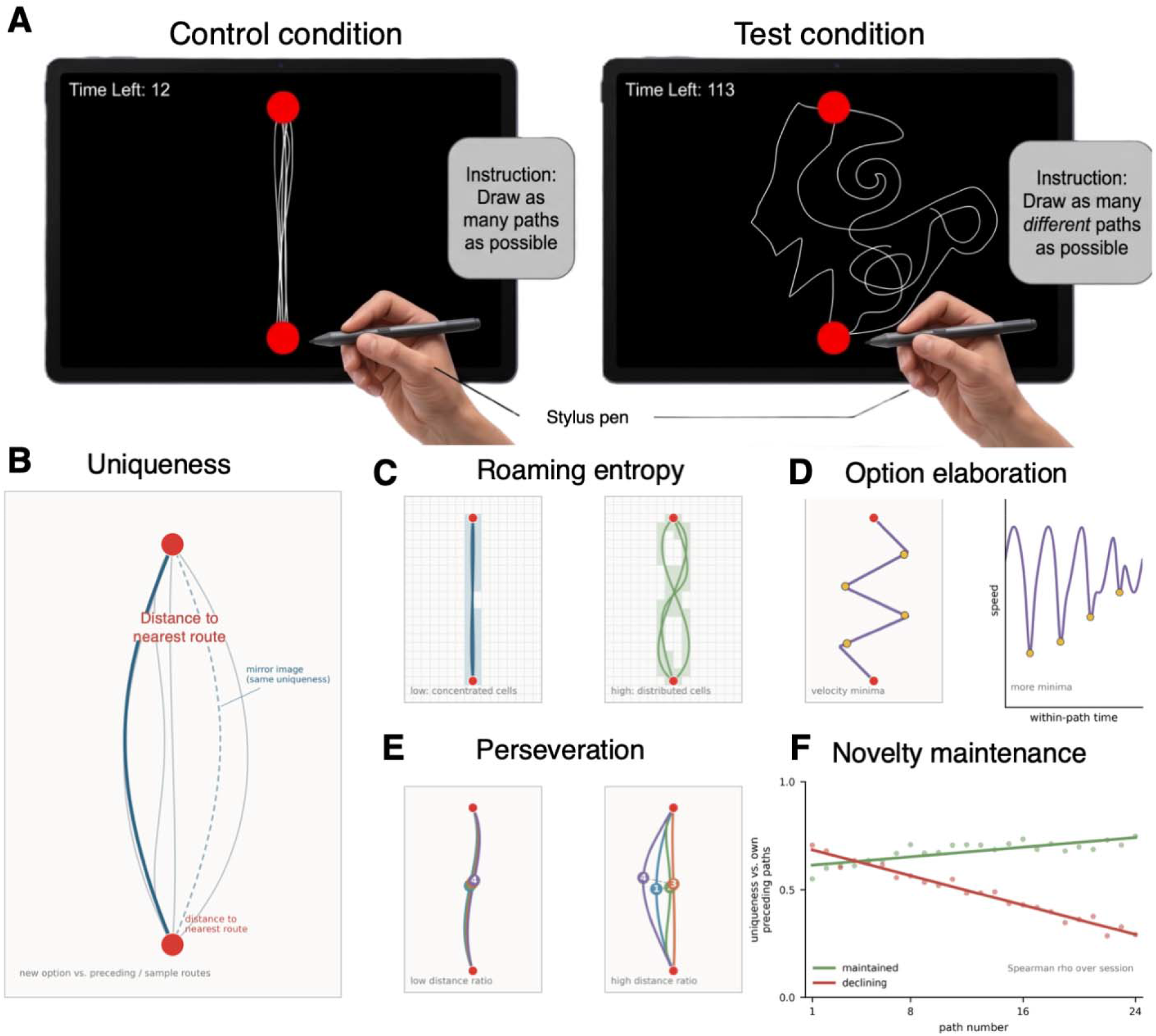
Option generation task and principal measures. (A) Option generation task illustration. In the control condition (left), participants drew as many paths as possible between two target circles (red) on a touchscreen tablet within 30 s. This condition measured baseline drawing speed. In the option generation (test) condition (right), participants drew as many *different* paths as possible between the same two circles within 120 s. Previously drawn paths remained visible on screen. (B) The primary outcome measure was path uniqueness, computed as the mean nearest-neighbour distance between each participant’s test routes and all other routes in the dataset, using a feature representation that incorporated position, velocity and acceleration profiles. Fluency (route count) and mean drawing speed were recorded in both conditions. (C) Roaming entropy was computed as the Shannon entropy of grid-cell visits across the search arena, normalised to lie in [0, 1]. (D) Option elaboration was computed as the within-path velocity minima (yellow dots), illustrated on a single trajectory (left) and on its corresponding speed-time profile (right). (E) Perseveration was computed as the ratio of mean consecutive-path distance to mean overall pairwise distance. Low values (left): successive paths trace near-identical trajectories. High values (right): consecutive paths are shape-distinct. Numbered paths show four ordered routes; dashed lines connect their midpoints. (F) Novelty maintenance was computed as the within-subject Spearman correlation between path order and prefix uniqueness (uniqueness of each path relative to one’s own preceding paths). Two illustrative time courses are shown: maintained novelty (green; stable or rising across the session) and declining novelty (red; falling across the session).

### Task data and principal measures

All behavioural preprocessing was performed using Python scripts. Route samples were grouped by task condition (control and test). Only completed paths were retained, as done previously^17^. For each path, the raw trajectory consisted of timestamped x-y coordinates. Participant-level summary metrics were computed separately for the control and test blocks, including route count and mean drawing speed.

The primary behavioural outcome was route uniqueness during the test block, computed following the approach described previously^17^. Each route was resampled to 200 equally spaced points along its arc length using linear interpolation. The resulting trajectory was converted into a feature vector comprising x position, y position, smoothed x and y velocity, and smoothed x and y acceleration. Velocity was computed as the first difference of position; acceleration as the first difference of velocity. Both were smoothed with a moving-average window of 40 samples and downsampled by a factor of 10 before concatenation with the full-resolution position features. To make the distance metric invariant to left-right mirroring, a horizontally flipped version of each route feature vector was also created (x-position, x-velocity and x-acceleration components negated).

Route uniqueness was computed using a nearest-neighbour approach across all test routes in the combined dataset (100 controls + 150 patients). That is, for each route, uniqueness was defined as the minimum Euclidean distance to any other test route in the full set, taking the smaller of the distance to the original and the mirrored version. Participant-level uniqueness was the mean of these route-wise minimum distances across all of participant’s test routes.

To examine whether differences in uniqueness could be explained by low-level motor features, we derived a motor control measure from the control condition. For each participant, control-condition routes were summarised by mean speed. We then computed a motor-residualised uniqueness measure by regressing raw mean uniqueness on mean control condition drawing speed, fitted across the pooled sample of controls and patients. Residualised uniqueness was defined as the residual from this regression. Similar analyses using other motor features from the control task, such as peak speed, path straightness (chord-to-arc ratio), tremor root-mean-square, or endpoint scatter and the coefficient of variation of path duration, all yielded similar results to mean speed. Mean speed in the control condition was therefore retained for simplicity.

To further test whether the group difference in uniqueness survived adjustment for demographic confounds, we fit a general linear model predicting raw uniqueness from control condition motor speed, age, sex and group (control vs. patient). Associations between motor speed, residualised uniqueness, and clinical variables were assessed using Spearman rank correlations, with 95% confidence intervals derived from 5,000 bootstrap resamples.

### Decomposition of uniqueness

To identify the behavioural mechanisms contributing to individual and group differences, as laid out in the CHANSS project aims^20^, we decomposed uniqueness into four complementary mechanistic features computed from the test block. First, ***roaming entropy*** quantified how broadly each participant sampled the spatial option space, using normalised Shannon entropy over occupied regions of a 16 × 24 cell grid discretising the task canvas. Higher values indicate more spatially distributed exploration. ***Option elaboration*** was indexed by the mean number of submovements per path, estimated from within-path velocity minima. This measure highly correlates with between-path deliberation time (see Results) and captures the degree of active within-path option development. ***Perseveration*** was measured as the mean distance between consecutive paths divided by the participant’s overall mean pairwise path distance, indexing whether successive options remained locally similar or moved through the option space. Lower values indicate more perseverative behaviour. ***Novelty maintenance*** was measured as the Spearman correlation between temporal position in the session (path number) and route uniqueness relative to one’s own preceding paths, capturing each participant’s ability to sustain unique option generation as the session progressed.

We quantified the contribution of each mechanism to residualised uniqueness using Shapley-value variance decomposition (Shapley R²). This approach decomposes total model R² by averaging each predictor’s marginal contribution over all possible subsets of the remaining predictors, yielding an exact additive partition of explained variance. All variables were standardised (z-scored) before entered into the regression analyses for the Shapley decomposition. We additionally assessed the degree to which the four mechanisms accounted for the patient-control uniqueness group difference through a stepwise attenuation analysis. Three ordinary least squares models were fit predicting raw uniqueness: (1) group alone; (2) group plus control condition motor speed; and (3) group plus motor speed plus the four mechanism measures. The percentage attenuation of the group coefficient was computed relative to the group-only model.

Finally, we examined whether patients could be separated into distinct cognitive-mechanistic subtypes using k-means clustering on the four standardised (z-scored within the patient sample) mechanism measures. We evaluated solutions from K = 1 to K = 6 and selected the number of clusters that maximised mean silhouette width. The final solution (K = 3) was fit with 500 random initialisations. Clusters were visualised using principal component analysis of the standardised mechanism space. To characterise the clinical relevance of the resulting clusters to the principal clinical measures associated with uniqueness, we compared residualised uniqueness, AMI behavioural subscale, BACS verbal fluency and CDS total score across clusters using non-parametric Kruskal–Wallis tests, with p-values adjusted for multiple comparisons using the Benjamini-Hochberg false discovery rate (FDR) procedure^31^.

Behavioural preprocessing and route-level uniqueness computation were performed in Python (version 3.11) using NumPy and Pandas. All statistical analyses were conducted in R (version 4.1).

## Results

### Sample characteristics and raw task performance

Patient demographic details are summarised in Table 1. Patients were overall older than the control group and included more males. Although performance in the task is invariant to gender and age^17^, we included these variables in the analyses below. In terms of self- and clinician-rated apathy scores, the patient population showed a large variability, with some participants scoring low in the no-apathy ranges while others score high in the severe apathy ranges^28,32^ (Fig. S1).

Raw measures of the task for controls and patients showed different patterns of results across control and test conditions (Fig. 2). In the control condition, healthy control participants generated more routes than patients (Fig. 2A, controls: M = 21.65, SD = 7.05, n = 100; patients: M = 15.36, SD = 8.82, n = 148; t(239.05) = 6.22, p < .001). Controls were also faster in their path drawing (Fig. 2B, controls: M = 1,959.65 px/s, SD = 662.30, n = 100; patients: M = 1,454.09 px/s, SD = 921.31, n = 148; t(245.03) = 5.03, p < .001). By contrast, in the test condition, route count did not differ between groups (Fig. 2C, controls: M = 53.50, SD = 25.49, n = 100; patients: M = 56.25, SD = 29.81, n = 150; t(233.08) = −0.78, p = 0.437). Similarly, mean speed did not differ between groups (Fig. 2D, controls: M = 1,280.15 px/s, SD = 542.39, n = 100; patients: M = 1,150.82 px/s, SD = 634.64, n = 150; t(233.15) = 1.72, p = 0.086).

**Figure 2.**
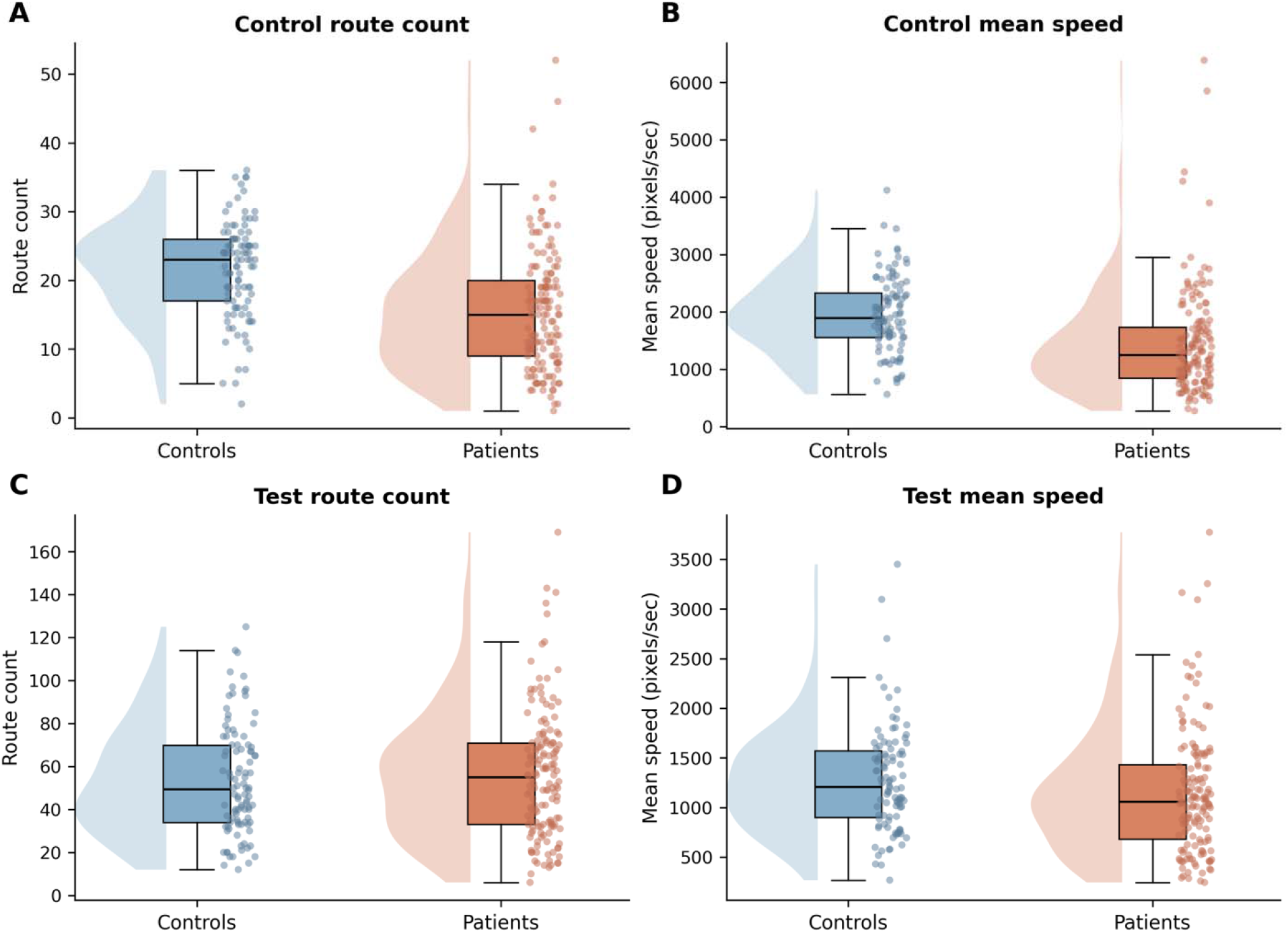
Behavioural performance in the control and test conditions. Raincloud plots (half-violin kernel density, box-and-whisker, jittered data points) show distributions for controls (blue) and patients with schizophrenia (orange). (A) Route count and (B) mean drawing speed in the 30 s control condition: patients produced fewer routes and were slower than controls. (C) Route count and (D) mean drawing speed in the 120 s test condition: route count did not differ between groups, while patients continued to draw more slowly.

### Patients generated less unique paths

The primary outcome, mean test uniqueness, was markedly reduced in patients compared with controls (Fig. 3A). Healthy control participants had double the mean uniqueness score observed in patients (controls: M = 682.52, SD = 446.69, n = 100; patients: M = 295.49, SD = 293.97, n = 150; *t*(155.80) = 7.63, p < .001). We tested whether the group difference in option generation uniqueness could be explained by more basic movement features, measured from stereotyped control task trajectories (see Methods). To this end, we residualised uniqueness by mean movement speed in the control task (Fig. 3B). Residualised uniqueness remained significantly smaller in patients compared to controls (controls: M = 206.28, SD = 453.95; patients: M = −139.38, SD = 292.73; *t*(154.25) = 6.73, p < .001). The results did not change when considering other low-level motor features, such as deviation from straight movements or high-frequency speed fluctuations.

**Figure 3.**
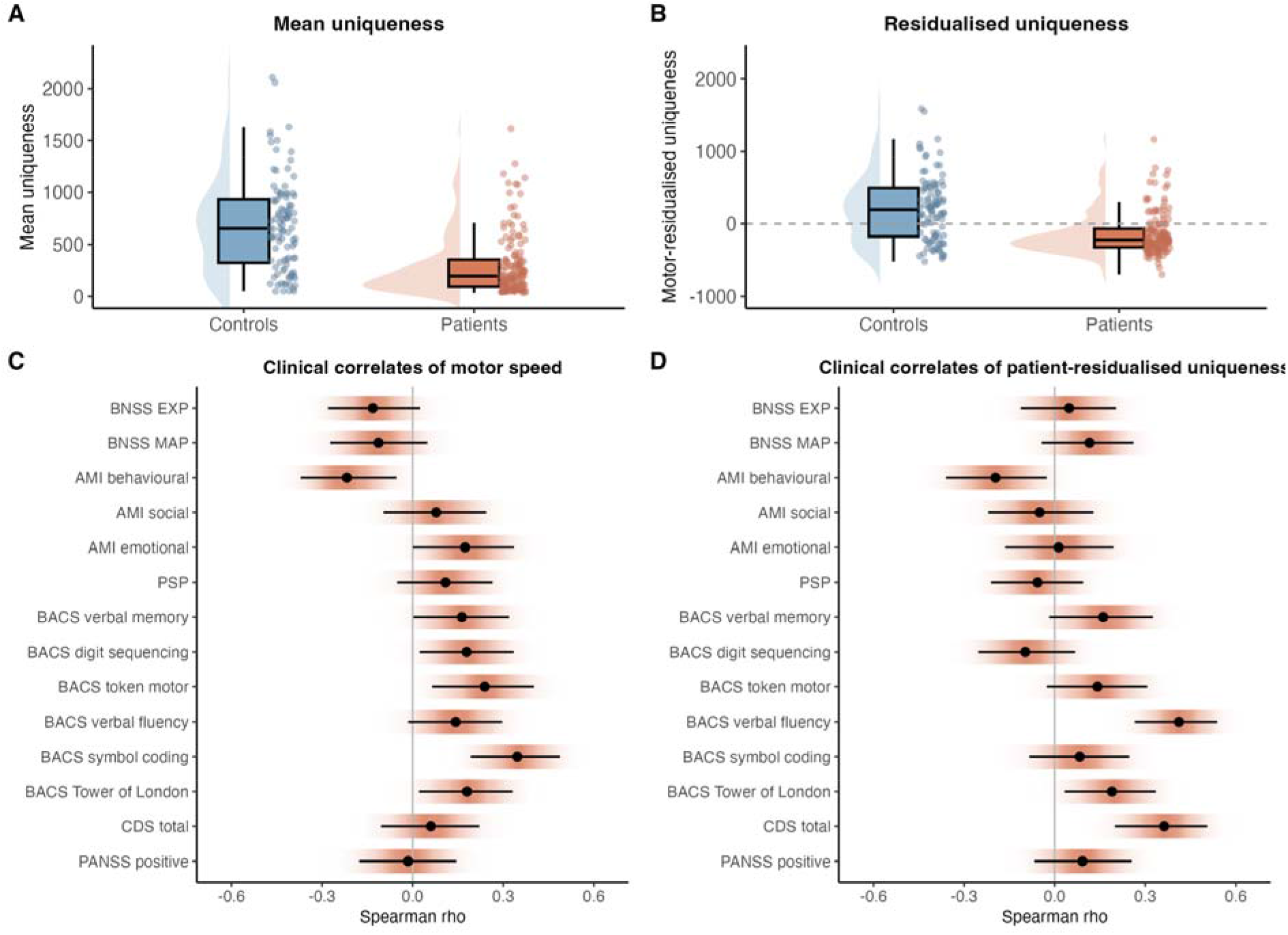
Principal outcome measures across groups and their associations with patient clinical variables. A) C) Associations between mean motor speed in the control condition and the different clinical variables. Spearman rho is shown, together with 95% confidence interval (error bars), computed by bootstrapping with 5000 samples with replacement (shaded density showing full bootstrap distribution space, using a kernel density estimation). D) Same as (C) but for motor-residualised uniqueness.

As control and patient group were different in age and sex, we further controlled and tested for the effect of these variables on uniqueness in a general linear model (Table 2). As previously reported, there was no effect of age on uniqueness^17^, and a non-reliable effect of sex. Importantly, the group difference remained significant when including these variables. Moreover, within the patient group, there was no effect of antipsychotic medication dose on uniqueness (Supplementary Material, Table S1)

**Table 2.** General linear model (GLM) examining demographic predictors of uniqueness. The GLM was significant (F(4, 231) = 23.94, p<0.001), with R^2^_adj_ = 0.28.

| Predictor | B estimate | SE | 95% CI | t statistic | p |
| --- | --- | --- | --- | --- | --- |
| Intercept | 706.07 | 98.43 | 512.14, 900.01 | 7.17 | $p < .001$ |
| Control<br>motor speed | 0.04 | 0.03 | -0.01, 0.09 | 1.42 | $p = 0.156$ |
| Sex (male) | 87.94 | 49.19 | -8.98, 184.86 | 1.79 | $p = 0.075$ |
| Age | -2.34 | 2.69 | -7.64, 2.96 | -0.87 | $p = 0.385$ |
| Group<br>(patients vs.<br>controls) | -421.34 | 68.14 | -555.59, -<br>287.08 | -6.18 | $p < .001$ |

We next examined the associations between the low-level motor features and main outcome measure of uniqueness (residualised for motor feature in control condition) with the principal cognitive variables (Fig. 3C-D). As expected, low-level motor speed in the control task was reliably associated with various cognitive domains in the BACS, namely motor tokens task (rho=0.24), symbol coding (rho=0.35) and Tower of London task (rho=0.18). Motor speed was also negatively associated with behavioural apathy, as measured by the AMI behavioural subscale (rho=-0.22).

By contrast, motor-residualised uniqueness was positively associated verbal fluency (rho=0.39) and Tower of London (rho=0.19). Clinically, uniqueness was positively associated with depressive symptom severity, as measured using the CDS (rho=0.36) and negatively associated AMI behavioural subscale (rho=-0.20). Together, the results suggest that, after taking individual differences in motor performance into account, reduced uniqueness in patients was associated with *increased* self-reported apathy but *reduced* depressive symptoms. We next examined the cognitive mechanisms underlying individual differences in (residualised) uniqueness.

### Reduced uniqueness in patients reflected distinct cognitive mechanisms

To identify cognitive mechanisms underlying reduced uniqueness in the patient group, we decomposed option generation behaviour into four complementary mechanistic features (see Figure 1C-1F). First, ***roaming entropy*** quantified how broadly each participant sampled the spatial option space, using normalised entropy over occupied regions of the task space. Second, ***option elaboration*** was indexed by the mean number of submovements per path; this measure was strongly correlated with test inter-path timing across participants (Spearman rho = 0.65), and captured within-path construction rather than pause duration. Third, ***perseveration*** was measured as the distance between consecutive paths relative to the participant’s overall pairwise path distances, indexing whether successive options remained locally similar or moved through the option space. Fourth, ***novelty maintenance*** was measured as the Spearman correlation between temporal position in the experiment (path number) and route uniqueness, capturing each participant’s ability to sustain unique option generation as the session progressed.

These four mechanism measures were weakly to moderately inter-correlated in patients (Fig. 4A; median absolute Spearman rho = 0.37; maximum absolute rho = 0.55), consistent with partially separable components of exploratory behaviour. Across the full sample, the four mechanisms explained 69.3% of the variance in motor-residualised uniqueness (Fig. 4B). Variance partitioning showed the largest contribution from roaming entropy (Shapley R^2^ = 0.287), followed by option elaboration (Shapley R^2^ = 0.208), perseveration (Shapley R^2^ = 0.156), and novelty maintenance (Shapley R^2^ = 0.041).

**Figure 4.**
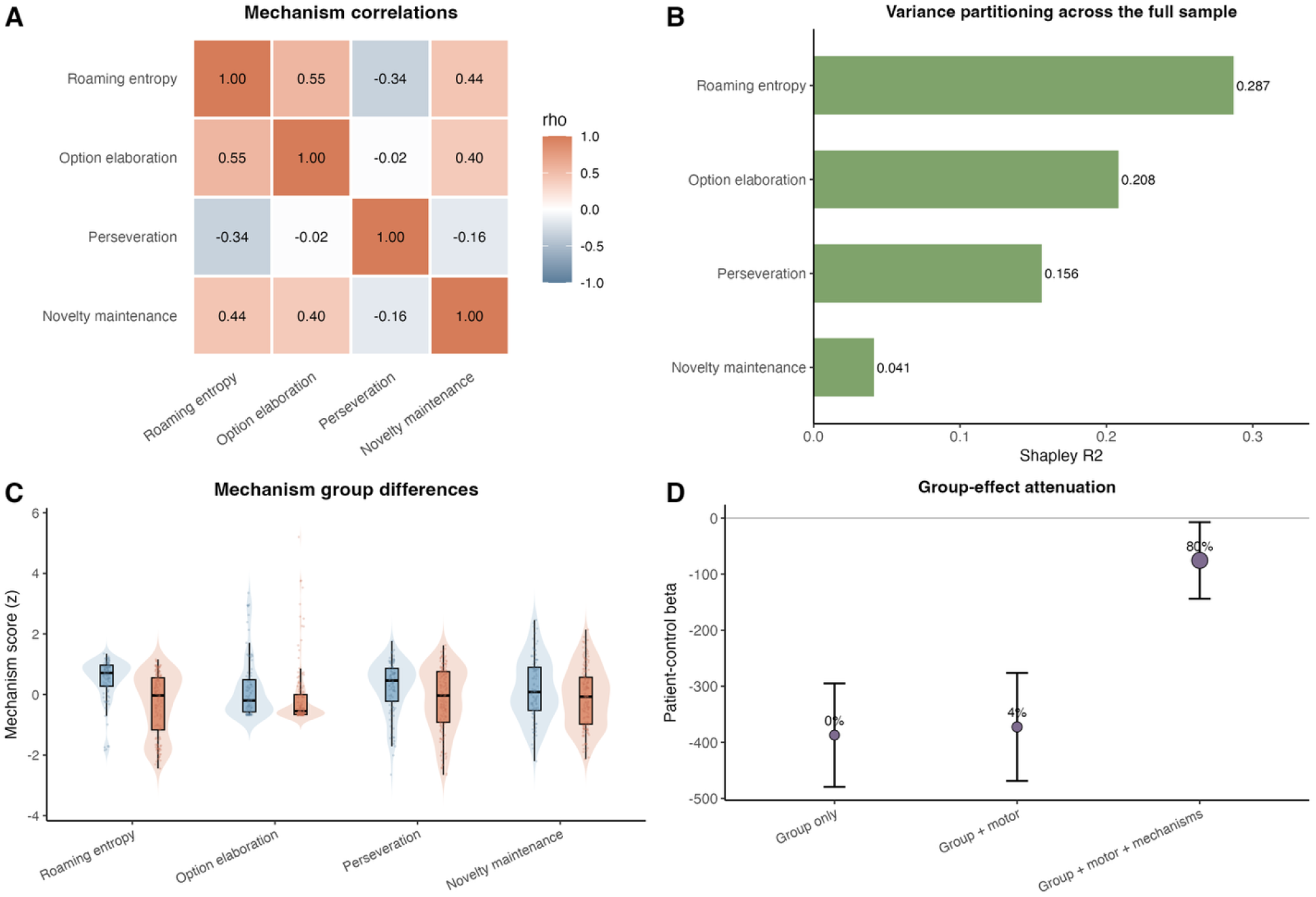
Exploratory cognitive mechanisms explaining uniqueness. (A) Spearman correlations among the four mechanism measures across all participants (patients and controls). Roaming entropy, option elaboration and novelty maintenance form a positively correlated cluster (ρ = 0.40 to 0.55); perseveration is weakly or negatively correlated with the others (ρ = −0.34 to −0.02). (B) Shapley R² decomposition of motor-residualised uniqueness across the full sample, jointly explaining ∼69% of variance. Roaming entropy is the dominant contributor, followed by option elaboration, perseveration, and novelty maintenance. (C) Raincloud plots of each mechanism (z-scored within the combined sample) for controls (blue) and patients (orange). Patients show reduced values across all four mechanisms. (D) Patient-control coefficient on uniqueness across nested OLS models (mean ± 95% CI). The unadjusted effect was minimally attenuated by motor covariates (4%) but reduced by 80% once the four mechanisms were added, indicating the patient-control gap is largely accounted for by cognitive mechanisms rather than basic motor capacity.

Compared with controls, patients showed significantly lower roaming entropy (0.768 vs 0.865; t(246.74) = 7.62, p < .001), reduced option elaboration (3.46 vs 5.23 submovements per path; t(211.09) = 2.24, p = .026), and lower perseveration ratio (0.776 vs 0.838; t(234.78) = 3.28, p = .001; Fig. 4C). Novelty maintenance was also reduced in patients, with a more negative temporal uniqueness association (−0.151 vs −0.064; t(202.75) = 2.00, p = .047), although the non-parametric comparison was weaker. Finally, adding motor speed alone produced little attenuation of the patient-control uniqueness effect (3.8%), whereas adding the four cognitive mechanisms (roaming entropy, option elaboration, perseveration and novelty maintenance) attenuated the group effect by 80.5% (Fig. 4D), indicating that reduced uniqueness in patients was largely captured by these distinct exploratory mechanisms. We next used these measures to examine different profiles of option generation behaviour in patients.

### Distinct cognitive profiles of apathy in schizophrenia

Finally, we asked whether patients could be separated into distinct cognitive-mechanistic profiles based on the four behavioural measures linked to uniqueness. We applied k-means clustering to the standardised mechanism measures in the patient group. Model selection supported a three-cluster solution, with K = 3 showing the highest silhouette width among the tested solutions (Fig. 5A; mean silhouette = 0.376). Demographic and clinical measures for the three clusters are reported in Table S1. The three clusters occupied partially separable regions of the patient mechanism space (Fig. 5B). Cluster profiles showed distinct scores across the four cognitive mechanisms (Fig. 5C). Cluster 1 was the smallest (n = 11), characterised by elevated scores across all four mechanism dimensions, particularly option elaboration. Cluster 2 was the largest subgroup (n = 83), showing relatively preserved roaming entropy, novelty maintenance and lower perseveration but little option elaboration. Cluster 3 (n = 56) showed the clearest exploratory impairment profile, with reduced roaming entropy, reduced novelty maintenance and relatively higher perseveration (Fig. 5C).

**Figure 5.**
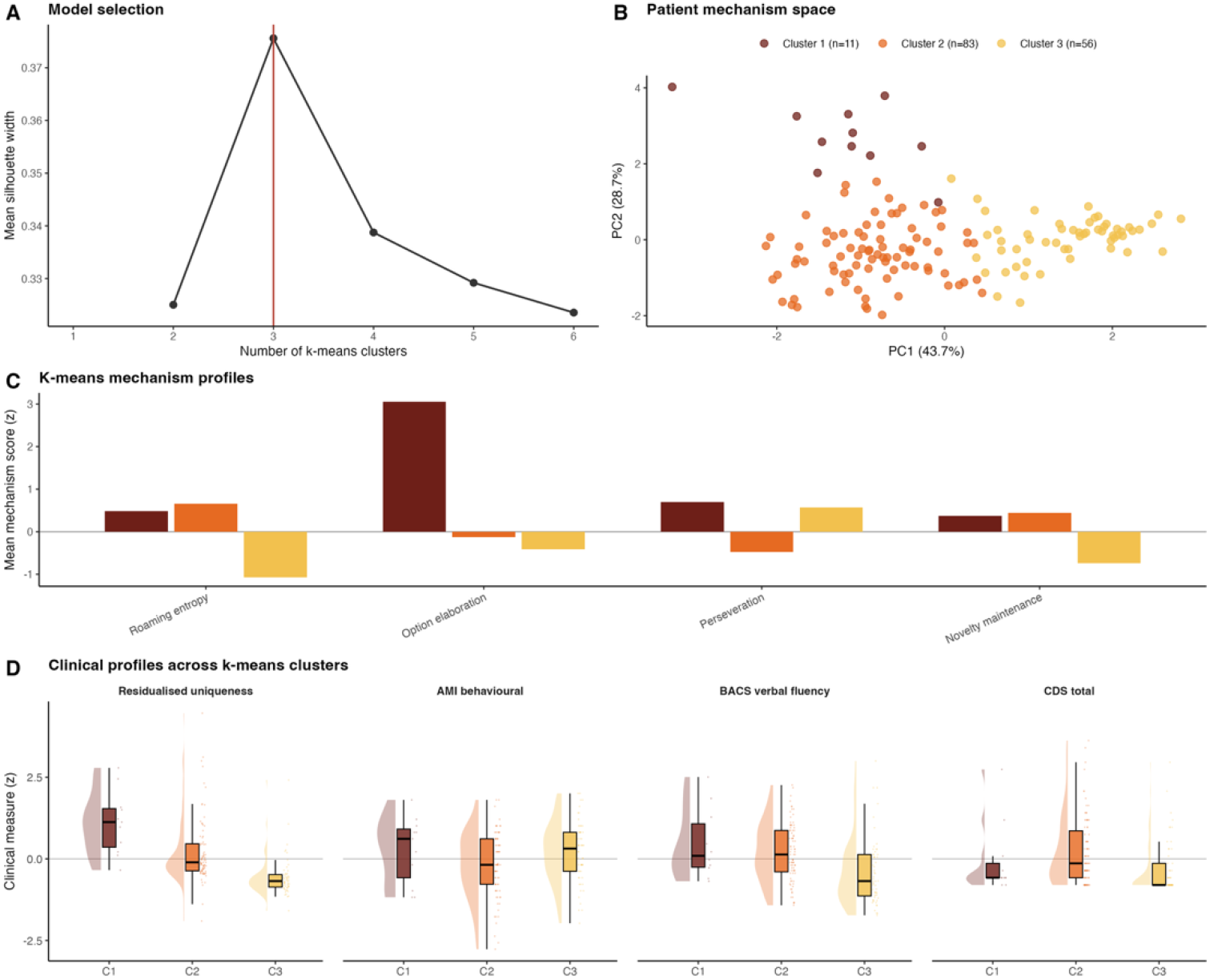
Profiles of apathy in the patient group. (A) Model selection: mean silhouette width across k = 2-6, peaking at k = 3 (vertical line). (B) Patient mechanism space projected onto the first two principal components (72.4% cumulative variance), with each patient coloured by k-means cluster assignment (Cluster 1, n = 11; Cluster 2, n = 83; Cluster 3, n = 56). (C) Cluster profile plot, showing mean standardised mechanism score within each k-means cluster. For each mechanism, values were z-scored across the patient clustering sample, then averaged separately within Cluster 1, Cluster 2, and Cluster 3. (D) Clinical profiles across clusters. Raincloud plots showing z-scored distributions of residualised uniqueness, AMI behavioural apathy, BACS verbal fluency, and CDS total depressive symptoms for each cluster.

These mechanism profiles were clinically meaningful (Fig. 5D). Residualised uniqueness differed strongly across clusters (Kruskal-Wallis H = 60.73, p < .001, q < .001), with the highest uniqueness in Cluster 1, intermediate values in Cluster 2, and the lowest uniqueness in Cluster 3. Clusters also differed in verbal fluency (H = 17.96, p < .001, q < .001), again with Cluster 3 showing the lowest performance. Depressive symptoms differed across clusters (CDS total: H = 17.89, p < .001, q < .001), with the highest scores in Cluster 2. Behavioural apathy was overall similar across the three clusters (AMI behavioural: H = 5.05, p = .080, uncorrected). Similarly, there were no significant differences in terms of antipsychotic doses (H = 0.21, p = 0.899, uncorrected). Thus, patient heterogeneity in option generation was not simply a severity gradient, as different mechanism profiles mapped onto distinct cognitive and clinical signatures.

## Discussion

This study investigated option generation in schizophrenia as a potential driver of reduced goal-directed behaviour and apathy. The large, multisite, multiethnic cohort supports the robustness and generalisability of this effect across different cultural and clinical settings, addressing a recognised need for greater population diversity in psychosis research^33^. The results reveal a marked deficit in the uniqueness, but not the quantity, of self-generated behavioural options. Patients produced options of roughly half the uniqueness of healthy controls. This deficit was not explained by fewer generated options and remained robust after adjustment for motor speed, age, and sex. Clinically, uniqueness was selectively associated with verbal fluency and self-reported behavioural apathy. It was also associated with higher depressive symptoms, which is consistent with the direction reported in major depression^24^. Mechanistic decomposition revealed four distinct processes contributing to reduced uniqueness, with reduced spatial roaming (exploration of the canvas) as the dominant contributor. K-means clustering revealed three distinct patient profiles defined by different mechanistic patterns and clinical correlates. Together, these findings illuminate distinct cognitive mechanisms underlying apathy in different individuals with schizophrenia, with implications for understanding heterogeneity in negative symptoms and for future treatment development and tailoring.

### Reduced option uniqueness in schizophrenia

The central finding of this study is that patients with schizophrenia generated options that were roughly half as unique as those of healthy controls, a deficit that was robust to adjustment for motor speed, age, and sex. Although a previous study examined the ability of schizophrenia patients to generate options verbally for actions in real-world scenarios^19^, it was not possible to quantify features of option generation related to a well-defined goal-directed behaviour as done in our study. Crucially, these features enabled us to show that the impairment in option generation was specific to the distinctiveness of generated options rather than their quantity, as route count (fluency) did not differ significantly between groups. The dissociation between preserved fluency and reduced uniqueness suggests that schizophrenia does not impair the capacity to initiate action per se, but rather the ability to diversify the content of self-generated behavioural options. Motor speed was not a significant predictor of uniqueness, confirming that the uniqueness deficit reflects a cognitive rather than motor limitation. Instead, motor speed was associated with distinct cognitive domains in the BACS, including token motor speed task, symbol coding, and Tower of London performance. By contrast, (motor-independent) uniqueness was selectively associated with verbal fluency cognitive subdomain, further supporting the cognitive link of the uniqueness finding.

This pattern extends previous work using the OGT. In healthy volunteers, dopaminergic manipulation modulated the fluency-uniqueness trade-off, with levodopa increasing route count at the expense of uniqueness^17^. Individuals with MDD produced fewer but more unique options than controls, a profile interpreted as reflecting reduced dopaminergic constraint on exploratory search^24^. The schizophrenia profile reported here is different, with preserved fluency but reduced uniqueness, and thus cannot be accounted for by the same mechanism. The results in schizophrenia are also different to findings in healthy individuals, wherein self-reported apathy was associated with lower fluency and, paradoxically, greater uniqueness, as also found in MDD^17,24^. That the two psychiatric conditions possibly most associated with apathy, schizophrenia and MDD, show opposite profiles on option uniqueness has theoretical and clinical implications to the mechanisms and potential treatment of apathy in these patient groups, which we discuss below.

### Clinical correlates: verbal fluency, apathy, and depression

The most robust cognitive correlate of uniqueness was verbal fluency as assessed by the BACS^25^. This association was substantially stronger than for any other cognitive subdomain. The selectivity of the verbal fluency association is notable: both the OGT and verbal fluency tasks require the spontaneous, sustained generation of outputs satisfying a certain constraint within a fixed time: phonological or semantic category in verbal fluency, and spatial distinctiveness in the OGT. The structural parallel suggests they share cognitive substrates.

In schizophrenia, verbal fluency deficits are among the most consistently replicated executive impairments and are closely linked to prefrontal dysfunction and negative symptoms^21,22^. Non-verbal fluency has been studied less extensively, but impairments have also been reported on figural fluency tasks^34^. Our results suggest that impaired option generation in schizophrenia may be another expression of this same executive-generation deficit, one that is culture-fair, motor-adjustable, and captures spatial rather than linguistic search. The advantage of the OGT over verbal fluency is that it does not depend on language, which is linked to crystallised intelligence and may be biased towards individuals with higher education^35^. By contrast, the main limitation of the OGT is that it depends on motor capacity; a limitation we addressed by measuring basic motor capacity in a control condition and accounting for it in the main analyses.

Alongside verbal fluency, depressive symptoms were positively associated with uniqueness, with higher uniqueness associated with higher depressive symptoms on the Calgary Depression Scale (rho = 0.35). The positive association between uniqueness and depressive symptoms in our schizophrenia sample reproduces the direction of findings reported in clinically depressed individuals^24^, suggesting that the depression-related uniqueness signature cuts across diagnostic boundaries.

In contrast to depressive symptoms, lower uniqueness was associated with greater self-reported behavioural apathy, and this relationship was stable under non-parametric bootstrapping. The magnitude of the association was modest, which may partly reflect the difficulty of capturing apathy constructs using self-report questionnaires^36^. The AMI which we used in our study was developed to address these challenges and to cover the three main constructs of apathy: emotional, social and behavioural, yet it was designed for the general population^28^. By contrast, existing self-report scales of negative symptoms in schizophrenia were designed to match the constructs assessed by clinician scales^37^, such as the BNSS^38^, rather than. Yet these scales for schizophrenia patients do not cover the neuroscience-informed constructs of apathy, as is possible with the AMI. Developing a new hybrid consensus scale for assessing self-report motivation in schizophrenia might address this gap.

In contrast to self-reported behavioural apathy, uniqueness was not correlated with clinician-rated negative symptom dimensions of MAP or EXP of the BNSS. This dissociation between self-rated apathy and clinician-rated negative symptoms echoes a recent study emphasising the poor alignment between patient- and clinician-reported apathy symptoms in schizophrenia^32^. Again, the lack of agreement in the constructs assessed between the different questionnaires^39^ might be profitably addressed in the development of new consensus scales^32^. Building on advances in neuroscience and biological understanding of motivation^40^, new scales are urgently needed to assess the experience of apathy in individuals with schizophrenia. Our study suggests that the development of new questionnaires can be further informed by clinically scalable neuroscience tasks^41^, which allow a better mechanistic understanding of complex mental health constructs.

### Mechanistic decomposition of the uniqueness deficit

Decomposing uniqueness into partially independent cognitive mechanisms, we identified reduced spatial roaming as the dominant contributor to the uniqueness deficit. Roaming entropy accounted for 41% of the explained variance in (motor-independent) uniqueness and showed the strongest group difference of all decomposed features. The degree of within-path complexity indexed by submovements, which we referred to as option elaboration, contributed a further 30% of explained variance, while perseveration and novelty maintenance made smaller but meaningful contributions. The four cognitive mechanisms (reduced spatial roaming, lower option elaboration, greater perseveration, and reduced novelty maintenance) together attenuated the patient-control uniqueness difference by over 80%, indicating that reduced uniqueness in patients was largely captured by these distinct exploratory processes rather than a unitary deficit.

This profile resembles what has been described as reduced exploration in foraging frameworks^42,43^: patients remained in familiar territories of the path space rather than venturing into novel regions. Reduced cognitive flexibility and restricted semantic search in verbal fluency tasks, whereby patients generate words from fewer semantic clusters and show reduced cluster-switching ^44,45^, may have a spatial analogue in the OGT. The dominant contribution of reduced roaming entropy in patients suggests a similar contraction of the behavioural search space: patients repeatedly traversed familiar regions of the option space without generating genuine alternatives. This has implications for understanding behavioural apathy in schizophrenia as arising, at least in part, from deficits in the cognitive capacity to conceive of alternative actions, rather than, or alongside, reduced valuation of those actions^18^.

While reduced action space has been suggested to lead to apathy by narrowing down the action repertoire^18^, how might depressive symptoms link (positively) to increased uniqueness? One possibility is that less spatially constrained search is a behavioural analogue of mind-wandering in depression^46–48^. Mind-wandering reflects reduced attentional anchoring to task-relevant goals, and it predisposes to the unconstrained, negatively valenced thought seen in rumination^49,50^. These hypotheses can be tested in future studies, for example by studying a potential link between state rumination and cognitive performance^51^ and option generation behaviour.

### Distinct patient profiles and implications for treatment

The k-means clustering analysis revealed that patients were not uniformly impaired in option generation but separated into three mechanistically distinct subtypes, similar in self-rated apathy scores. The smallest cluster (n = 11) showed a healthy-like profile with preserved or elevated scores across all four OGT task dimensions, and correspondingly high uniqueness. The largest cluster (n = 83) showed preserved spatial exploration with high roaming entropy, maintained novelty, and low perseveration, but importantly showed reduced option elaboration and the highest depressive symptom scores. The third cluster (n = 56) showed the most pronounced exploratory impairment, with markedly reduced roaming entropy, reduced novelty maintenance, and elevated perseveration; this subgroup also had the lowest verbal fluency and the lowest uniqueness scores.

These profiles are clinically meaningful in several respects. First, they demonstrate that the overall group-level uniqueness deficit is not evenly distributed across patients but is driven disproportionately by the impaired exploration subgroup. Second, the dissociation between clusters supports the independence of the underlying mechanisms: preserved spatial roaming coexisted with elevated depressive symptoms, while impaired roaming was accompanied by reduced verbal fluency rather than depression. This pattern is consistent with the opposing associations between uniqueness and apathy versus depression observed at the full sample level and suggests that these associations may partly reflect distinct patient subgroups rather than competing influences within the same individuals.

The clinical profiles may point to two distinct subgroups. In the first, uniqueness is reduced because patients explore the action space less (reduced roaming entropy). In the second, patients explore the space adequately but, because of low mood, lack the motivation to develop each course of action beyond a stereotyped pattern. This echoes with a potential separation between primary and secondary negative symptoms—that is, motivational problems occurring as part of the core schizophrenia deficit vs. those motivational problems resulting from related features of the condition, such as depression^52,53^ and defeatist beliefs ^54,55^. Similarly, these findings are consistent with the notion that seemingly similar cognitive impairments can result from reduced motivation and inflated effort perception or alteration to the cognitive apparatus carrying out the cognitive task itself^40^. Interestingly, the small cluster showing comparable uniqueness to that of healthy controls suggests relatively intact option generation in that subgroup. Yet the similar levels of self-rated apathy in the ‘healthy-like’ cluster points to a different mechanism of apathy, possibly ‘upstream’ to an executive motivation dysfunction, e.g., in decision-making or metacognition; a hypothesis that will be explored in future studies of the CHANSS project^20^.

The existence of mechanistically separable profiles has implications for treatment. If reduced spatial exploration identifies a subgroup with executive generative impairments linked to verbal fluency, interventions targeting cognitive flexibility, such as cognitive remediation approaches^56^ focusing on set-shifting and divergent thinking and creativity^57^, may be particularly effective for this subgroup. Conversely, the subgroup with preserved exploration but elevated depression and reduced development of novel options may benefit more from interventions targeting mood and motivational engagement, such as behavioural activation^58,59^. More broadly, the OGT may serve as a brief, repeatable behavioural assay for stratifying patients according to the cognitive structure of their motivational impairment, complementing traditional symptom-based classification.

### Limitations and future directions

Several limitations should be noted. First, the present analyses are cross-sectional, precluding causal inference. Longitudinal work will be needed to determine test-retest reliability and whether option generation impairment predicts functional outcome. Second, The CHANSS sample spans a wide range of illness stages and recruitment sites across three countries, which is a strength for generalisability but introduces heterogeneity. The OGT’s culture-fair, non-linguistic format makes it well suited to cross-cultural application, as confirmed by the multilingual task implementation (English, Spanish, Mandarin). Third, although the four-mechanism decomposition explained a substantial proportion of variance in uniqueness (69%), the mechanisms were derived from the same task data as the primary outcome and were largely exploratory and were based on previous literature. Independent validation using external measures of spatial exploration, cognitive flexibility, and perseveration would strengthen the claim that these reflect dissociable cognitive processes. Fourth, the k-means clustering, while revealing interpretable structure, was applied to a moderately sized sample and should be replicated in larger cohorts.

## Conclusion

Schizophrenia is associated with a selective impairment in the uniqueness of behavioural option generation: patients draw as many paths as controls but generate far less unique ones, a deficit driven primarily by reduced spatial roaming and reduced within-path elaboration. This impairment is not reducible to motor slowing, is selectively associated with verbal fluency and self-rated behavioural apathy, and is directionally opposite to the uniqueness profile observed in depression. Mechanistically distinct patient profiles map onto different clinical signatures, suggesting that heterogeneity in option generation reflects separable cognitive pathways to apathy in schizophrenia. These findings establish option generation as a clinically relevant, mechanistically tractable behavioural phenotype in schizophrenia, distinct from standard neurocognitive measures, and suggest that apathy in schizophrenia may partly reflect an impoverished repertoire of self-generated action options due to distinct cognitive and motivational impairments.

## Data Availability

All data produced in the present study are available upon reasonable request to the authors

## Acknowledgements

We thank Dr Yuen Ang for sharing his analysis code with us.

## Supplementary Material

**Figure S1.**
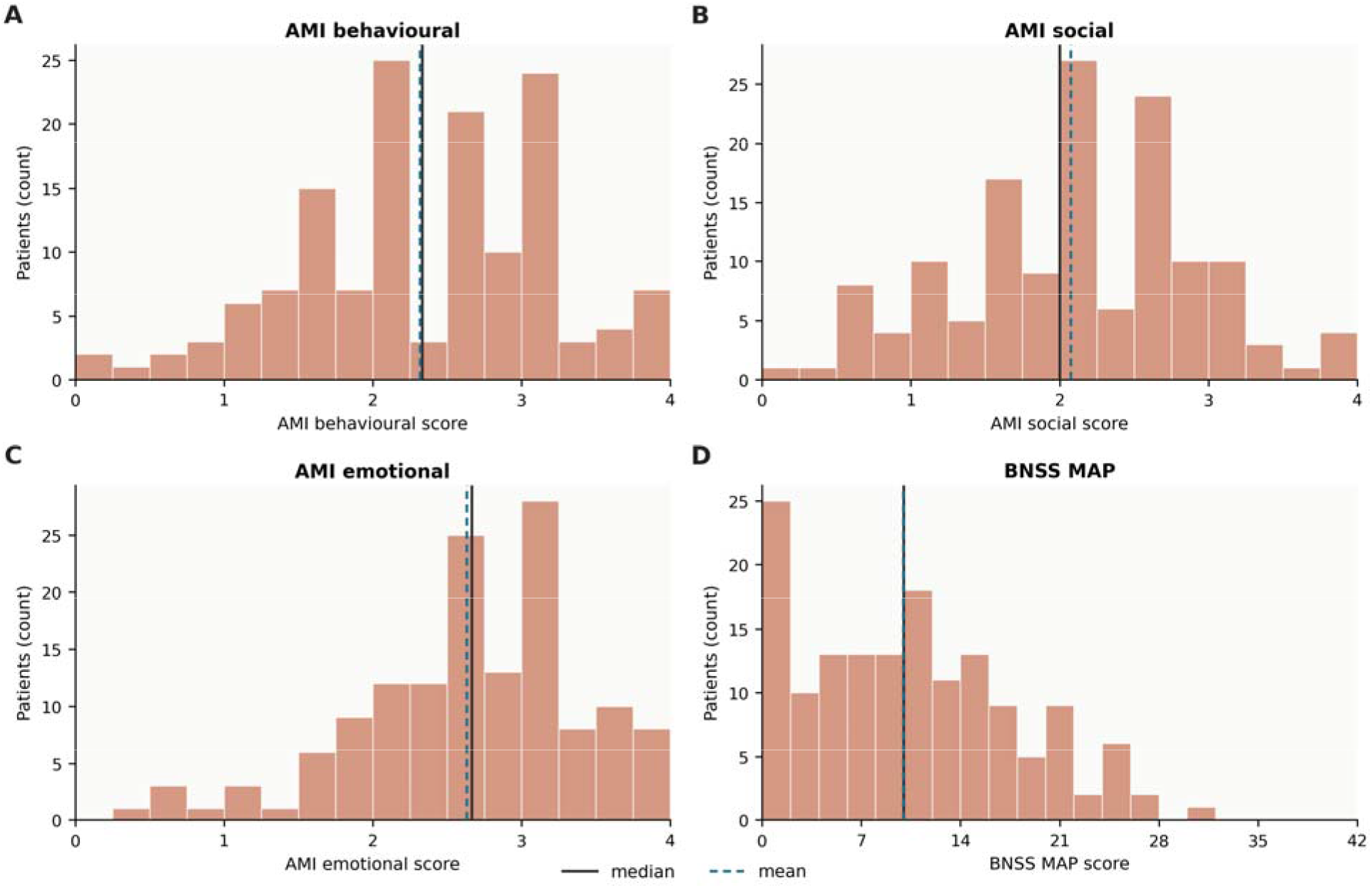
Patient self- and clinician-reported apathy. Patients showed a large variability in self- (A-C) and clinician-rated apathy scores. (A) Distribution of apathy motivation index (AMI) behavioural subscale, across patients in the study. Mean score was 2.31 which is considered moderate apathy. (B) Same as (A), but for social AMI. Mean score was 2.075 which is considered below the moderate apathy cut-off. (C) Same as A-B but for AMI emotional subscale. Mean score was 2.63 which is considered severe apathy. (D) Distribution of the clinician rated Brief Negative Symptom Scale (BNSS) Motivation and Pleasure (MAP) subscale. Mean score was 9.95 which is considered below the cut-off for significant apathy. Cut-offs were based on

---

**Table S1.** Antipsychotic effect on uniqueness. General linear model within the patient group, predicting uniqueness from control condition motor speed, sex, age, and antipsychotic burden expressed as percentage of British National Formulary maximum dose.

| Predictor | B estimate | SE | 95% CI | t statistic | p |
| --- | --- | --- | --- | --- | --- |
| Intercept | 269.03 | 118.91 | [33.96, 504.09] | 2.26 | p = 0.025 |
| Control motor speed | 0.05 | 0.02 | [0.00, 0.10] | 2.05 | p = 0.042 |
| Sex (male) | 5.00 | 49.07 | [-92.00, 102.00] | 0.10 | p = 0.919 |
| Age | -2.30 | 2.36 | [-6.97, 2.36] | -0.98 | p = 0.331 |
| Antipsychotic burden | 0.54 | 0.51 | [-0.47, 1.54] | 1.06 | p = 0.290 |

